# Residency Program Directors’ Selection Criteria: An Analysis of the NRMP Match from 2008-2024

**DOI:** 10.64898/2026.11.22.26368534

**Authors:** Dhruva Patel, Allison Dalton, Steven Roth

## Abstract

**Purpose:** COVID-19 shifted in-person to virtual residency interviews beginning in the 2020– 2021 application cycle, raising concerns that program directors (PDs) would rely more heavily on objective metrics to compensate for challenges in assessing personal characteristics. Whether this transition altered the underlying ranking criteria, and whether such changes persisted, has not been systematically evaluated.

**Method:** Data were extracted from the NRMP Biennial Survey for Anesthesiology (ANES), Internal Medicine (IM), General Surgery (GS), and Radiology (RAD) from 2008–2024. The authors collected PD-reported mean importance ratings (1–5) and the percentage citing each factor across five thematic domains, focusing on the top three factors/theme and items cited important by ≥ 50% of PDs. Trends were analyzed using generalized estimating equations with wild cluster bootstrap and Benjamini-Hochberg correction.

**Results:** Mean importance ratings increased significantly over time, with slopes 0.039 (ANES) to 0.071 (IM) points per survey cycle, which survived bootstrap and Benjamini-Hochberg correction in both the 3-item and ≥ 50%-cited models except ≥ 50%-cited Radiology. With only 2-3 panel units per theme, no theme-by-specialty trend met the corrected threshold across the 20 tests (all q ≥.17). In a post hoc pooled interview-day model, importance rose by 0.061 points per survey cycle (95% CI 0.052-0.069), with no reliable evidence that slopes differed by specialty (wild bootstrap p =.07) and a downward level shift at the 2021 survey wave (-0.26 points, 95% CI-0.32 to-0.20).

**Conclusions:** Interview-day importance rose over time across all four specialties, with a downward deviation in the 2021 survey wave rather than a steady rise, but no individual theme-by-specialty trend met the corrected threshold. Although these findings cannot establish whether programs changed how they used interview time, we can conclude it maintains high importance in the ranking process despite widespread change to a virtual interview process.

**Graphical Abstract:** 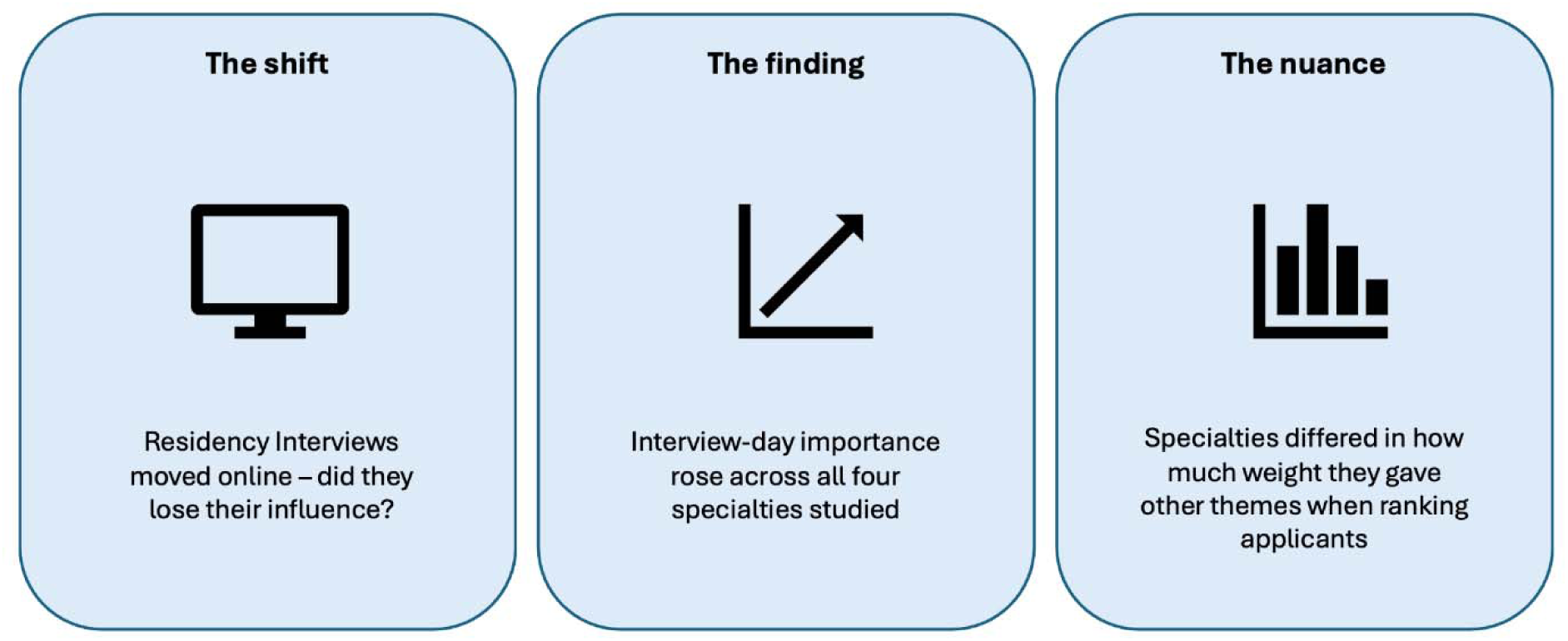

**Teaser Text:** A study analyzing the National Resident Matching Program data from 2008-2024 found that interview-day importance rose over time, with a deviation coincident with the 2021 shift to virtual residency interviews.

There is a Methods Supplement and Supplemental tables

---

Accurate and consistent ranking criteria are central to the success of the residency match, shaping not only the outcomes, but also how well applicants fit their programs’ clinical, academic, and cultural environments. The COVID-19 pandemic created a major shift from in-person to virtual interviews beginning in 2020–2021.^1,2^ Program directors (PD) across specialties found it significantly more difficult to assess applicants’ interpersonal skills, professionalism, and programmatic “fit” in the virtual format.^3^ All interviews had previously been conducted in person. Programs adopted novel recruitment strategies to compensate for loss of face-to-face interaction, including a second visit and a pre-interview social with current residents.^1,4^

Specialty-specific literature published since the transition suggests these challenges were widely shared but not uniformly addressed.^3,5,6^ Previous commentary anticipated that General Surgery (GS) programs would place greater weight on objective data, such as standardized letters of recommendation and skills testing, to stratify an applicant pool they could no longer evaluate in person. PDs broadly reported greater difficulty assessing fit, personality, and communication skills virtually based on single-program survey data.^6,7^ Some programs have switched to hybrid interviews, i.e., offering applicants the option for a virtual or in-person interview, whereas other specialties, like Internal Medicine (IM), remained virtual-only after the pandemic.^7,8^ Similarly, Orthopedic Surgery PDs uniformly described altering their selection processes, placing greater emphasis on communication from faculty mentors, direct emails from applicants, home-applicant status, and attendance at virtual information sessions amid a sharp rise in application volume.^5^ Most of these studies are limited as single institution studies and have not included large numbers of programs or candidates.

The virtual interview era prompted shifts in how applicants were evaluated, yet whether these shifts extended to the underlying criteria PDs use to rank applicants, and whether they persisted or represented a lasting change to the selection process, remains unclear. From 2008-2024, the number of residency programs participating in the National Resident Matching Program (NRMP) increased from 4,064 to 6,395 while applicants increased from 28,737 to 44,853.^9,10^

The NRMP surveys PDs biennially about factors influencing applicant ranking. Data include percentage of PDs endorsing importance of a specific factor and mean importance scores. There were five themes in the survey. These repeated, standardized measures offer a large scale data approachto examine impact of virtual interviewing on the factors valued by PDs, across specialties and over time.

We focused on Anesthesiology (ANES), GS, IM, and Radiology (RAD) because they represent a mix of competitiveness tiers, rely on a comparable standardized application/rating system, and showed divergent post-pandemic interview policies (hybrid vs. virtual-only). This cross-specialty design allowed us to test whether shifts in applicant evaluation were a specialty-specific response or part of a broader trend across residency selection since these four specialties accounted for 34% of active residents across ACGME-accredited specialties and subspecialties in 2024–2025.^11,12^ This study aimed to test the hypothesis that residency programs changed their areas of emphasis and weighted scoring when reviewing applicants due to inability to conduct in-person interviews. We studied longitudinal trends in program factor ratings by examining changes in average importance scores and percentage cited across application cycles before and after the COVID-19–associated transition to virtual interviewing.

## Methods

The study received exempt status from the Institutional Review Board of the University of Illinois at Chicago. Data were extracted from the NRMP’s PD Survey reports for ANES, IM, GS, and RAD from 2008-2024. We accessed the NRMP website in May 2026 and captured in Excel (Microsoft 365 Enterprise, Redmond, Washington, USA) the PD-reported mean importance ratings (1–5, where 5 indicated the highest importance) and the percentage of PDs citing each factor when constructing their rank order lists. The five themes, 1) education and performance, 2) interview day, 3) other knowledge of applicants, 4) personal qualities, and 5) standard licensure test, comprised 15, 7, 7, 13, and 10 items, respectively. Survey years were 2008, 2010, 2012, 2014, 2016, 2018, 2020, 2021, 2022, and 2024, but 2022 was excluded because it listed only 5 factors, yielding nine repeated cross-sectional measurements over time. For a baseline, 2014 was used due to the report in years prior not stating percentage of PDs citing each factors’ importance.

Percentage of PDs citing a factor and mean importance rating was collected for the top three ranking factors of the five themes as categorized in the 2024 report for all survey years (3-item analysis). As an alternate test, we examined items in the five themes cited as important by ≥ 50% of PDs. Because the percentage of PDs citing each factor is first reported for 2014, the ≥ 50%-cited models covered the six survey years 2014, 2016, 2018, 2020, 2021, and 2024. The ≥ 50% threshold was applied separately to each specialty in each survey year, so factors could enter or leave across years and the panels are unbalanced. Selecting the 3-item factors from the 2024 report conditions a longitudinal analysis on its endpoint. The 3-item factors, their wording, their availability by year and specialty, and their percentage citing are listed in **Supplementary Table 1,** and the factors entering the ≥ 50%-cited models are listed in **Supplementary Tables 2 and 3**.

Analysis was conducted with Stata v19 (Stata, College Station, TX, USA) using Generalized Estimating Equations (GEE) with a Gaussian family and identity link, treating each selection factor as the repeated panel unit across survey years and specifying an exchangeable working correlation structure. The wild cluster bootstrap was implemented with boottest.

Due to the explicit data and criteria labeling, blinding of the authors was not feasible. Theme-level models were estimated first, and any factors within statistically significant themes (p <.05) were examined individually using multivariable regression and Bonferroni-corrected pairwise year comparisons. To identify the years of most rapid change in perceived importance for each selection factor, a Gaussian curve was fit to the longitudinal ratings, and its derivative (dydx, i.e., marginal effect) was used to locate the inflection points where the rate of change in importance was greatest.

For theme 5, standard licensure test, cluster-robust ordinary least squares (OLS) was used in place of GEE because the “any failed USMLE attempt” item appeared only from 2016 onward, and the resulting unbalanced panel prevented stable estimation of the exchangeable working correlation matrix. That item was therefore excluded, and the remaining items were modeled by OLS with standard errors clustered on selection factor.

Because each theme’s panel comprised only 2-3 selection factors, cluster-robust and GEE sandwich standard errors at the theme level are unreliable on their own. We therefore also assessed theme-level p values with a wild cluster bootstrap-t (9,999 replications, Webb weights, clustered on selection factor).^13^ The Benjamini-Hochberg procedure was applied to the bootstrap p values to control the false discovery rate across the 20 theme-by-specialty tests. Bootstrap p-value floors, tests for nonlinearity, the segmented model for a level and slope change at the 2021 transition, and leave-one-survey-cycle-out sensitivity influence analyses are described in **Supplemental Methods**. Because the bootstrap does not eliminate the uncertainty arising from only 2-3 clusters per theme, theme-level findings are treated as exploratory.

The number of selection factors (clusters) and observations in every model is reported in **Supplementary Table 4**.

## Results

Across all four specialties, mean importance ratings of applicant selection factors were similar at baseline (ANES: 4.20 [0.38]; GS: 4.21 [0.36]; IM: 4.16 [0.38]; RAD: 4.24 [0.40]; mean [SD], **Table 1**) and increased significantly over the study period when all five themes were modeled jointly. This upward trend held in every specialty, although the rate of increase varied: average marginal effect (dydx) of survey year ranged from 0.039 (ANES) to 0.071 (IM), with IM showing the steepest rate of change, Radiology the second steepest (0.051), and Anesthesiology the most modest (0.039). Baseline estimates with 95% confidence intervals (CI), and marginal effects with SD and 95% CI, are in **Table 1**. Both the three-item and ≥ 50% importance ratings evaluations showed statistically significant results on naive testing. Applying a wild cluster bootstrap and Benjamini-Hochberg correction, the three-item model’s trend remained statistically significant after correction in all four specialties (bootstrap p =.004; Benjamini-Hochberg q ≤.007). The ≥ 50% model’s trend remained significant after correction in three of four (ANES, GS, IM; q ≤.011) but not RAD (naive p =.017, bootstrap p =.20, q =.20).

**Table 1:** Comparison of trend estimates across all five themes: full (3-item per theme) vs ≥ 50%-cited item analyses, by specialty.

| **Specialty** | **Baseline (2014) mean (SD) [95% CI]** | **3-item model** |  |  | **≥ 50%-cited model** |  |  |
|---|---|---|---|---|---|---|---|
|  |  | **dydx (SD) [95% CI]<sup>a</sup>** | **Naive p** | **Bootstrap p (q)** | **dydx (SD) [95% CI]<sup>a</sup>** | **Naive p** | **Bootstrap p (q)** |
| ANES | 4.20 (0.38) [4.07, 4.34] | 0.039 (0.040) [0.018, 0.059] | < .001 | .003 (.006) | 0.013 (0.023) [0.004, 0.022] | .005 | .006 (.008) |
| GS | 4.21 (0.36) [4.08, 4.35] | 0.046 (0.042) [0.025, 0.067] | < .001 | .004 (.006) | 0.015 (0.023) [0.005, 0.024] | .002 | .009 (.010) |
| IM | 4.16 (0.38) [4.04, 4.29] | 0.071 (0.053) [0.045, 0.098] | < .001 | < .001 (< .001) | 0.020 (0.032) [0.008, 0.031] | .001 | < .001 (< .001) |
| RAD | 4.24 (0.40) [4.12, 4.37] | 0.051 (0.038) [0.032, 0.070] | < .001 | < .001 (< .001) | 0.011 (0.023) [0.002, 0.019] | .017 | .20 (.20) |

Comparison of trend estimates across all 5 themes collapsed into one: full (3-item per theme) and ≥ 50%-cited item analyses, by specialty. Estimates are from GEE models; p values are shown for naïve and wild cluster bootstrap-p values (Webb weights, 9,999 replications, clustered on selection factor; 0 reported as <.001) with Benjamini-Hochberg q values across all eight specialty-by-model tests in parentheses). Average marginal effects reflect the change in mean importance rating per unit time (per survey for the 3-item model; per calendar year for the ≥ 50%-cited model; The 3-item model reflects the top-3 items per theme; the ≥ 50%-cited model was restricted to items cited as important by ≥ 50% of program directors. ANES = anesthesiology; CI = confidence interval; dydx = average marginal effect of survey year; GEE = generalized estimating equations; GS = general surgery; IM = internal medicine; RAD = radiology; SD = standard deviation.

Notes: **a.** Time units differ between models: the 3-item model’s dydx is per survey; the ≥ 50%-cited model’s dydx is per calendar year. Magnitude comparisons between columns should be interpreted as directional rather than strictly proportional.

On unadjusted testing, and bootstrap (**Table 2**), the Interview Day importance trends over time were significant in all four specialties (except for bootstrap p for Anesthesiology). Unadjusted testing was significant for all specialties for importance of Standard Licensure Test. The trends in importance of the other themes were significant over time in only some specialties: Education & Performance (IM, RAD), Other Knowledge of Applicant (ANES, IM, RAD), and Personal Qualities (ANES, GS). Because each theme-by-specialty model was estimated from only 2-3 panel units, below the number required for reliable cluster-robust or GEE sandwich inference, we reassessed these p values by wild cluster bootstrap with Benjamini-Hochberg correction across the 20 theme-by-specialty tests. No theme-level trend met the corrected threshold (all q ≥.17; **Table 2**), including Interview Day, for which bootstrap p values of.046-.057 in GS, IM, and RAD all corrected to q =.174. Correction applied to naive rather than bootstrap p values would have yielded q =.02 for Radiology, overstating the evidence; the Radiology interview-day trend is not distinguishable from the other three specialties at the theme level.

**Table 2:**
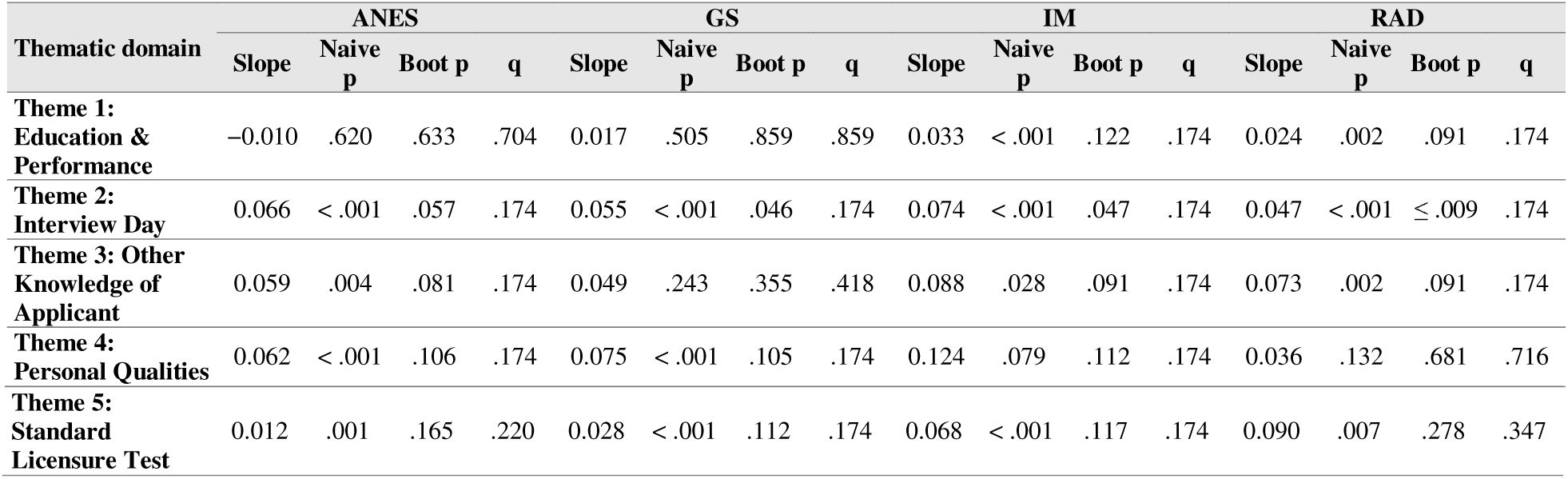
Comparison of thematic trends by specialty.

Because the smallest attainable bootstrap p with 3 clusters is limited, we fit a post hoc pooled interview-day importance model across specialties (12 factors, 108 factor-year observations; factor fixed effects; standard errors clustered on factor within specialty; **Supplementary Table 3 and 5**). Importance rose by 0.061 points per survey (95% CI 0.052-0.069; wild bootstrap 95% CI 0.052-0.069, p <.001). A Wald test of specialty-specific slopes was significant with cluster-robust inference (F[3, 11] = 7.76, p =.005) but not under the wild cluster bootstrap (p =.070), so we do not interpret specialty differences as established.

The interview-day importance trend was not well described by a linear slope: a quadratic term was significant in all four specialties (p =.001-.02), and a segmented model identified a downward deviation at the 2021 survey wave (joint p =.001-.017 on cluster-robust testing, interpretable only descriptively with 3 clusters) superimposed on an overall upward trend that kept the same direction when the trend was re-estimated after omitting each survey cycle in turn (**Figure 1**). In the pooled segmented model, the level change in 2021 was-0.26 points (95% CI - 0.32 to-0.20) and the change in slope was-0.06 points per survey (95% CI-0.15 to 0.02; p =.14); only two surveys (2021 and 2024) follow the transition, so these estimates are imprecise. Segmented estimates for each theme-by-specialty model, with 95% confidence intervals, are in **Supplementary Table 3**. This deviation is coincident with the 2021 survey wave; the repeated cross-sectional design has no comparison group unaffected by the pandemic, so it cannot be attributed to virtual interviewing.

**Figure 1.**
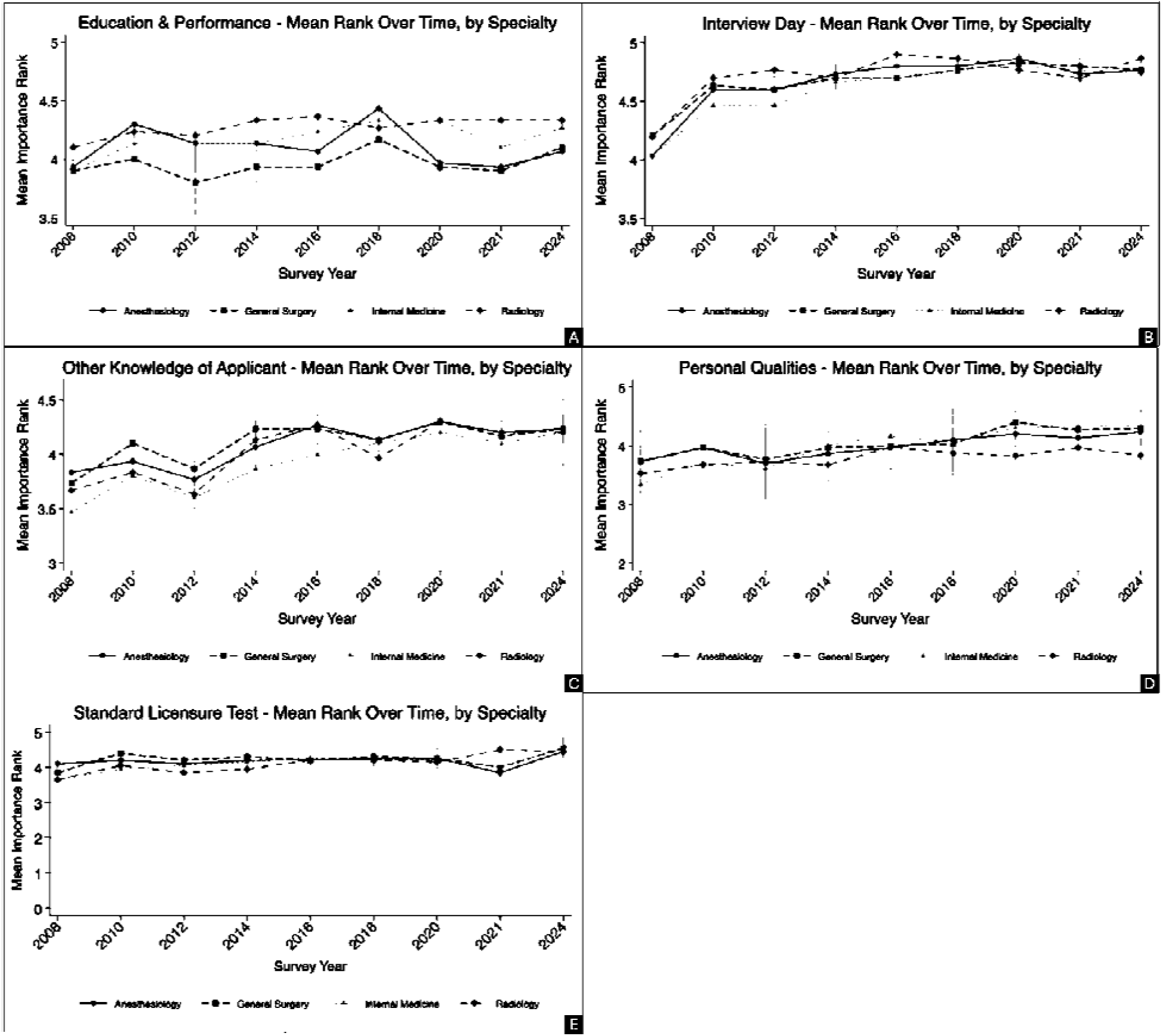
Mean importance rating of factors within all 5 thematic domains by specialty and survey year, 2008–2024 for top 3-items. Points are model-derived marginal means (GEE for Themes 1–4; OLS for Theme 5); error bars represent 95% confidence intervals. Graph A represents education and performance. B represents interview day Graph C represents other knowledge of applicant. Graph D represents personal qualities. Graph E represents standard licensure test.

Theme composites showed moderate raw shared variance (r = 0.13-0.75; VIF 1.3-3.3) that largely reflected common time trends and baseline differences between specialties: after removing specialty and linear year effects, residual correlations fell to r = 0.25-0.51 and VIFs to 1.4-1.6, below conventional thresholds for collinearity (**Supplemental Table 6; Supplemental Methods)**.

Comparison of thematic trends by specialty in each of the five themes. Cell entries are naive p values from a continuous year-sequence slope test within each thematic domain for the top 3 items (GEE; Standard Licensure Test modeled via cluster-robust ordinary least squares excluding the “any failed USMLE attempt” item); values <.05 indicate a statistically significant trend on unadjusted testing. Values in parentheses are the wild cluster bootstrap-p value (Webb weights, 9,999 replications, clustered on selection factor) and the Benjamini-Hochberg q value across the 20 theme-by-specialty tests, calculated on the bootstrap p values. With 3 clusters (2 for Standard Licensure Test) the smallest attainable bootstrap p is approximately.009 (.056); the Radiology Interview Day bootstrap p is reported at this floor (raw bootstrap p <.001). ANES = anesthesiology; GEE = generalized estimating equations; GS = general surgery; IM = internal medicine; OLS = ordinary least squares; RAD = radiology. The identity and number of factors (clusters) and observations in each model is in **Supplementary Table 1**

When restricting to factors cited by ≥ 50% of program directors, the aggregate upward trend replicated across all four specialties (**Table 1**); baseline (2014) mean importance ratings were similar across specialties (range: 4.16–4.24; **Supplemental Table 7**), and IM retained the steepest slope in both analyses. No theme-by-specialty trend survived Benjamini-Hochberg correction of the bootstrap p values (minimum q =.30, Personal Qualities in ANES; **Supplemental Table 7**); naive p values as small as <.001 corresponded to bootstrap p ≈.11, consistent with the anti-conservative bias of cluster-robust standard errors with as few as 2-10 panel units. Theme-level findings from this restricted analysis are therefore not interpreted as robust.

In contrast, a post hoc pooled interview-day model for the ≥ 50%-cited items (16 factors, 96 factor-year observations; **Supplemental Table 7**) showed a positive trend of 0.005 points per calendar year (95% CI 0.001-0.009; wild bootstrap 95% CI 0.0015-0.0084, p =.007), with no evidence that slopes differed by specialty (bootstrap p =.46) and a level shift at 2021 of-0.09 points (95% CI-0.14 to-0.05). Item-level results were broadly consistent with these theme-level patterns, with leadership qualities (ANES), feedback from current residents (GS), perceived interest in program (IM), personal prior knowledge of the applicant (RAD), and USMLE Step 1 pass status (RAD) each emerging as significant drivers within their respective themes on naive, uncorrected testing.

## Discussion

This study examined whether the shift to virtual interviewing altered the factors residency PDs use to rank applicants, using longitudinal NRMP data over a 16-year period and across four diverse specialties: anesthesiology, general surgery, radiology, and internal medicine, that constitute about 1/3 of all of the participants in the NRMP. Because no established standard exists for evaluating PD rankings of factors deemed important, we used two models: a top 3-item model and a ≥ 50%-cited model. Both showed the same significant upward trend, although their slopes are expressed in different time units and so are not directly comparable in magnitude. Overall importance ratings for selection criteria increased significantly across all four specialties over the study period, with “interview day” performance emerging as the most consistent contributor to this upward trend, suggesting that interview-day importance rose over the study period, including around the 2021 survey wave, rather than falling.

This finding runs counter to the assumption motivating our original hypothesis that losing face-to-face interaction would incentivize PDs to rely more heavily on objective, non-interview metrics (e.g., standardized test scores, letters of recommendation, and education performance) to compensate for a diminished ability to assess interpersonal qualities remotely.^14^ An explanation, which these data cannot test, is that programs adapted their interview process, for example, adding additional means for applicants to connect with programs through informal conversations, or resident-led sessions, rather than abandoning the interviews’ importance altogether. This interpretation is consistent with prior reports of programs developing novel recruitment strategies to compensate for loss of in-person contact.^15^ Another possibility is that interviews retain some selection value in a virtual format because applicants’ verbal communication and highly visible nonverbal behaviors remain observable. A meta-analysis of 63 studies of employment selection found that nonverbal cues such as professional appearance, eye contact, and head movement were associated with the ratings of interview performance, and that these associations were largely unaffected by interview modality.^16^ If translatable to the residency program interviews, there may be key universally expected behaviors that, if not present, render the candidate less likely to be highly ranked.

We observed meaningful differences between specialties in the specific factors PDs valued during the interview period. On unadjusted testing, ANES and GS showed stronger upward trends in ratings of interpersonal skills, faculty interactions, and house staff feedback, while IM showed a stronger upward trend in educational performance ratings. These differences may reflect underlying differences in program culture and workflow; anesthesiology and surgery are often described as prioritizing team dynamics and interpersonal compatibility, given the close, high-stakes and intense nature of clinical collaboration between faculty and trainees, whereas internal medicine’s broader case mix and larger resident cohorts may favor more standardized, quantifiable measures of academic performance.^17–19^ These specialty-level patterns in the Personal Qualities and Education & Performance themes did not survive wild cluster bootstrap and Benjamini-Hochberg correction for the small number of panel units and multiple testing and therefore should be treated as hypothesis-generating rather than confirmed. Interestingly, within ANES, no single itemized factor rose to statistical significance on its own; rather, an overall sense of interpersonal fit appeared to be the more consistent driver of rank position. This suggests that some of the most influential information PDs use may not map cleanly onto the discrete categories NRMP surveys ask PDs to rate.

Our findings also add nuance to concerns raised in the literature about the limitations of virtual interviewing. In prior studies, faculty noted that virtual formats may allow applicants to present a curated version of themselves during short, structured interactions, whereas unscripted, informal moments, such as dinners or social events, were more likely to reveal an applicant’s “true” interpersonal qualities.^14^ Despite this concern, our data did not show PDs compensating by shifting weight toward other, more “objective” factors, which may indicate either that PDs remain confident in their ability to extract meaningful signal from brief virtual interactions, or that other unmeasured adaptations (e.g., changes to interview structure, number of interviewers, or informal virtual social events) are absorbing this concern in ways not captured by the NRMP survey categories.

Several limitations should be considered. The number of independent panel units (selection factors) per theme was small, 2 to 3, with approximately 30-50 typically needed for cluster-robust or GEE sandwich inference to be reliable. We addressed this limitation with wild cluster bootstrap and Benjamini-Hochberg correction, after which no theme-level finding met the corrected threshold; the smallest attainable bootstrap p (about.009 with 3 clusters and.056 with 2) limits what these tests can detect. The pooled interview-day models did not model correlation among factors from different specialties surveyed in the same year, so their standard errors may be understated. Item composition also changed within some themes, so trends partly reflect changes in which factors were surveyed rather than change within a constant item set (**Supplementary Table 3**). The concurrent shift of USMLE Step 1 to pass/fail scoring in 2022 overlaps with our study period and may have confounded interpretation of trends in standardized test performance. As students increasingly complete Step 2 earlier in their training to provide programs with more complete scoring information, later cycles in our dataset may not yet reflect the full downstream impact. Furthermore, item sets differed partially across specialties (e.g., items assessing personal statement content and leadership qualities were not surveyed in all specialties), which precluded a single unified item-level factor analysis. Additionally, our reliance on NRMP-defined factor categories may obscure more holistic or qualitative judgments that PDs report relying on but that do not correspond neatly to a single survey item or theme such as candidate-reported activities and research.^20^ Prevailing definitions of professionalism and desirable applicant qualities are themselves evolving, and it remains uncertain how the shift to virtual interviewing has influenced PDs’ evaluation of these qualities, particularly as norms around what constitutes appropriate applicant conduct continue to change.^21^

Our analysis was limited to four specialties with different interview structures or resident-selection cultures that may show different patterns, and future work should expand this comparison. The broader move toward more holistic review criteria parallels similar shifts in medical school admissions and may plausibly support efforts to recruit a more diverse applicant pool; however, our data do not capture the racial or other demographic characteristics of applicants or resident classes, and we are therefore unable to assess whether these changes in selection criteria have had any measurable effect on the diversity of matched residents.^22^ Finally, as a survey-based dataset, NRMP data reflect PDs’ self-reported priorities rather than observed ranking behavior, which may not fully align.

Taken together, these findings suggest that the transition to virtual interviewing was not accompanied by a decline in the importance PDs place on the interview itself. Whether programs changed how they use that opportunity, and what they listen for, is a hypothesis these data cannot test. Future research using hybrid interview data and more granular measures of interpersonal assessment may help clarify how programs are adapting their evaluation processes in the post-pandemic era.

## Conclusion

In a pooled model, interview-day importance rose over time across ANES, GS, IM, and RAD, with a downward deviation at the 2021 survey rather than a steady rise. Other apparent shifts in selection emphasis did not meet the corrected threshold given the small number of panel units and multiple comparisons, are exploratory, and warrant confirmation in larger, item-level datasets.

## Data availability

NRMP Program Director Survey reports are publicly available, and the complete extracted dataset is available at UIC Indigo (provide web link).

## Ethical approval

The study was deemed “exempt” as non-human research by the Institutional Review Board of the University of Illinois at Chicago

## Use of Artificial Intelligence

Statistical code for the GEE models was developed with assistance from Claude (Anthropic), and Claude assisted in manuscript editing.

## Previous presentations

Abstract was presented at the Great Lakes Anesthesia Meeting, August 28-30, 2026, Chicago, IL.

## Data availability

NRMP Program Director Survey reports are publicly available and the complete extracted dataset is available at UIC Indigo (link here)

## Data Availability

All data produced in the present study are available upon reasonable request to the authors

