## Supplemental Methods and Tables for "Residency Program Directors’ Selection Criteria: An Analysis of the NRMP Match from 2008-2024"

**Wild cluster bootstrap and p-value floors:**

Theme-level p values were reassessed with a wild cluster bootstrap-t using Webb six-point weights and 9,999 replications, clustered on selection factor, with the null imposed. With G clusters the Webb distribution contains only 6^G weight vectors (36 for G = 2 and 216 for G = 3), equivalent to 18 and 108 distinct absolute t statistics respectively. The smallest attainable bootstrap p value is therefore approximately .056 when a theme comprised two selection factors and .009 when it comprised three, irrespective of effect size. Bootstrap p values at these floors are reported as such; the floor, not the data, is the binding constraint for those cells. Cluster and observation counts for every model are given in **Supplementary Table 2**.

**Multiplicity:**

The Benjamini-Hochberg procedure was applied to the bootstrap (not naive) p values to control the false discovery rate at .05 across the 20 theme-by-specialty tests, separately within the 3-item and the ≥ 50%-cited analyses. Applying the same procedure to naive p values yields materially smaller q values and overstates the evidence, as noted in the Results.

**Functional form and segmented models:**

Departure from a linear trend was assessed by adding a quadratic term in survey sequence and by Ramsey RESET on the fitted values. Segmented models added an indicator for survey waves from 2021 onward (level change) and an interaction between that indicator and centered survey sequence (slope change), tested jointly by Wald test. Because only two survey waves (2021 and 2024) follow the transition, post-transition slope estimates are imprecise and the level change should be read as a descriptive deviation at the 2021 wave rather than as an effect of virtual interviewing; the repeated cross-sectional design provides no comparison group unaffected by the pandemic.

**Influence:**

Leave-one-survey-cycle-out analyses re-estimated each trend after omitting one survey cycle at a time (nine re-estimations per 3-item model, six per ≥ 50%-cited model). The sign and approximate magnitude of the pooled interview-day slope were preserved in every re-estimation.

**Theme composites and collinearity:**

To describe overlap among themes, a composite was formed for each theme as the mean rating of that theme's items within each specialty-year (n = 36 specialty-year observations; Theme 5 excluded the “any failed USMLE attempt” item). The variance inflation factor of each composite was obtained from its regression on the other four composites. No multivariable outcome model using the composites as predictors was fit, so these values describe overlap among the thematic domains rather than collinearity in an estimated model. Raw pairwise correlations ranged from r = 0.13 to r = 0.75 (VIF 1.3-3.3); Interview Day, Other Knowledge of Applicant, Personal Qualities, and Standard Licensure Test showed moderate-to-high shared variance (r = 0.57-0.75), whereas Education & Performance was comparatively distinct (r = 0.13-0.41). Repeating the calculation on residuals from a regression of each composite on specialty and survey year reduced correlations to r = 0.25-0.51 and VIFs to 1.4-1.6 (**Supplementary Table 6**), indicating that much of the raw overlap is attributable to shared time trends and baseline differences between specialties. This descriptive analysis cannot exclude other sources of overlap.

Supplemental Table entries are naive (uncorrected) p values, as in **Table 2.** When the same wild cluster bootstrap and Benjamini-Hochberg correction applied to **Table 2** was applied to these 20 ≥5 0%-restricted theme-by-specialty tests, no cell survived correction (minimum q = .30); several of the smallest naive p values shown (e.g., Radiology Standard Licensure Test and Other Knowledge of Applicant, both p < .001) corresponded to bootstrap p values of only .10-.11, reflecting anti-conservative bias with as few as 2-3 panel units rather than a genuine trend. These values should therefore be interpreted as exploratory. Factors and observations in each ≥ 50% model are given in **Supplementary Table 2.**

**Supplementary Table 1. Inventory of the 60 selection factors in the 3-item analysis (top three factors per theme, 4 specialties × 5 themes), with availability by survey year.**

| **Spec.** | **Theme** | **Factor (wording as extracted; spelling and capitalization harmonized)** | **Mean rating available (survey years)** | **% citing available (survey years)** | **% citing, 2014 → 2024** | **Years factors were ranked ≥ 50% (n/N)** | **≥ 50% in 2024** |
| --- | --- | --- | --- | --- | --- | --- | --- |
| ANES | Education & Performance | MSPE | All 9 | All 6 | 74 → 67 | 6/6 | Yes |
| ANES | Education & Performance | Class ranking/quartile | All 9 | All 6 | 75 → 50 | 5/6 | Yes |
| ANES | Education & Performance | Grades required in clerkships | All 9 | All 6 | 59 → 46 | 4/6 | No |
| ANES | Interview Day | Interpersonal skills | All 9 | All 6 | 91 → 90 | 6/6 | Yes |
| ANES | Interview Day | Interaction with faculty during interview and visit | All 9 | All 6 | 96 → 85 | 6/6 | Yes |
| ANES | Interview Day | Feedback from current residents | All 9 | All 6 | 81 → 69 | 6/6 | Yes |
| ANES | Other Knowledge of Applicant | Personal prior knowledge of the applicant | All 9 | All 6 | 66 → 52 | 6/6 | Yes |
| ANES | Other Knowledge of Applicant | Letters of recommendation | All 9 | All 6 | 65 → 64 | 6/6 | Yes |
| ANES | Other Knowledge of Applicant | Audition elective/rotation within your department | All 9 | All 6 | 68 → 51 | 5/6 | Yes |
| ANES | Personal Qualities | Personal statement | All 9 | All 6 | 60 → 64 | 5/6 | Yes |
| ANES | Personal Qualities | Leadership qualities | All 9 | All 6 | 75 → 64 | 6/6 | Yes |
| ANES | Personal Qualities | Perceived commitment to specialty | All 9 | All 6 | 72 → 62 | 5/6 | Yes |
| ANES | Standard Licensure Test | USMLE Step 2 | All 9 | All 6 | 69 → 74 | 6/6 | Yes |
| ANES | Standard Licensure Test | USMLE Step 1 pass | All 9 | All 6 | 88 → 60 | 6/6 | Yes |
| ANES | Standard Licensure Test | Any failed attempt in USMLE | 2016–2024 (5) | 2016–2024 (5) | — → 47 | 1/5 | No |
| GS | Education & Performance | MSPE | All 9 | All 6 | 58 → 61 | 5/6 | Yes |
| GS | Education & Performance | Class ranking/quartile | All 9 | All 6 | 55 → 53 | 5/6 | Yes |
| GS | Education & Performance | Grades required in clerkships | All 9 | All 6 | 41 → 49 | 2/6 | No |
| GS | Interview Day | Interpersonal skills | All 9 | All 6 | 89 → 93 | 6/6 | Yes |
| GS | Interview Day | Interaction with faculty during interview and visit | All 9 | All 6 | 91 → 88 | 6/6 | Yes |
| GS | Interview Day | Feedback from current residents | All 9 | All 6 | 76 → 88 | 6/6 | Yes |
| GS | Other Knowledge of Applicant | Personal prior knowledge of the applicant | All 9 | All 6 | 53 → 64 | 6/6 | Yes |
| GS | Other Knowledge of Applicant | Letters of recommendation | All 9 | All 6 | 79 → 69 | 6/6 | Yes |
| GS | Other Knowledge of Applicant | Audition elective/rotation within your department | All 9 | All 6 | 48 → 49 | 1/6 | No |
| GS | Personal Qualities | Personal statement | All 9 | All 6 | 51 → 63 | 5/6 | Yes |
| GS | Personal Qualities | Leadership qualities | All 9 | All 6 | 59 → 63 | 6/6 | Yes |
| GS | Personal Qualities | Perceived commitment to specialty | All 9 | All 6 | 69 → 64 | 6/6 | Yes |
| GS | Standard Licensure Test | USMLE Step 2 | All 9 | All 6 | 79 → 63 | 6/6 | Yes |
| GS | Standard Licensure Test | USMLE Step 1 pass | All 9 | All 6 | 82 → 49 | 5/6 | No |
| GS | Standard Licensure Test | Any failed attempt in USMLE | 2016–2024 (5) | 2016–2024 (5) | — → 36 | 0/5 | No |
| IM | Education & Performance | MSPE | All 9 | All 6 | 79 → 75 | 6/6 | Yes |
| IM | Education & Performance | Class ranking/quartile | All 9 | All 6 | 71 → 60 | 6/6 | Yes |
| IM | Education & Performance | Grades required in clerkships | All 9 | All 6 | 55 → 49 | 4/6 | No |
| IM | Interview Day | Interpersonal skills | All 9 | All 6 | 95 → 88 | 6/6 | Yes |
| IM | Interview Day | Interaction with faculty during interview and visit | All 9 | All 6 | 92 → 84 | 6/6 | Yes |
| IM | Interview Day | Interaction with house staff during interview and visit | All 9 | All 6 | 82 → 66 | 6/6 | Yes |
| IM | Other Knowledge of Applicant | Personal prior knowledge of the applicant | All 9 | All 6 | 59 → 35 | 4/6 | No |
| IM | Other Knowledge of Applicant | Letters of recommendation | All 9 | All 6 | 65 → 61 | 6/6 | Yes |
| IM | Other Knowledge of Applicant | Audition elective/rotation within your department | All 9 | All 6 | 47 → 35 | 1/6 | No |
| IM | Personal Qualities | Diversity characteristics | 2021–2024 (2) | 2021–2024 (2) | — → 61 | 2/2 | Yes |
| IM | Personal Qualities | Evidence of professionalism and ethics | 2010–2024 (8) | All 6 | 64 → 64 | 6/6 | Yes |
| IM | Personal Qualities | History of overcoming significant obstacles | All 9 | All 6 | 36 → 61 | 3/6 | Yes |
| IM | Standard Licensure Test | USMLE Step 2 | All 9 | All 6 | 86 → 78 | 6/6 | Yes |
| IM | Standard Licensure Test | USMLE Step 1 pass | All 9 | All 6 | 83 → 68 | 6/6 | Yes |
| IM | Standard Licensure Test | Any failed attempt in USMLE | 2016–2024 (5) | 2016–2024 (5) | — → 64 | 5/5 | Yes |
| RAD | Education & Performance | MSPE | All 9 | All 6 | 78 → 70 | 6/6 | Yes |
| RAD | Education & Performance | Class ranking/quartile | All 9 | All 6 | 73 → 77 | 6/6 | Yes |
| RAD | Education & Performance | Grades required in clerkships | All 9 | All 6 | 59 → 57 | 6/6 | Yes |
| RAD | Interview Day | Interpersonal skills | All 9 | All 6 | 97 → 87 | 6/6 | Yes |
| RAD | Interview Day | Interaction with faculty during interview and visit | All 9 | All 6 | 95 → 87 | 6/6 | Yes |
| RAD | Interview Day | Interaction with house staff during interview and visit | All 9 | All 6 | 85 → 80 | 6/6 | Yes |
| RAD | Other Knowledge of Applicant | Personal prior knowledge of the applicant | All 9 | All 6 | 55 → 53 | 5/6 | Yes |
| RAD | Other Knowledge of Applicant | Letters of recommendation | All 9 | All 6 | 65 → 43 | 5/6 | No |
| RAD | Other Knowledge of Applicant | Audition elective/rotation within your department | All 9 | All 6 | 50 → 43 | 2/6 | No |
| RAD | Personal Qualities | Personal statement | All 9 | All 6 | 49 → 57 | 5/6 | Yes |
| RAD | Personal Qualities | Perceived commitment to specialty | All 9 | All 6 | 61 → 57 | 6/6 | Yes |
| RAD | Personal Qualities | History of overcoming significant obstacles | 2021–2024 (2) | 2021–2024 (2) | — → 57 | 2/2 | Yes |
| RAD | Standard Licensure Test | USMLE Step 2 | All 9 | All 6 | 53 → 80 | 6/6 | Yes |
| RAD | Standard Licensure Test | USMLE Step 1 pass | All 9 | All 6 | 85 → 50 | 6/6 | Yes |
| RAD | Standard Licensure Test | Any failed attempt in USMLE | 2016–2024 (5) | 2016–2024 (5) | — → 33 | 1/5 | No |

Supplementary Table 1: The 3-item analysis retained three factors per theme per specialty, defined from the 2024 NRMP report; the full item pools per theme are larger (15, 7, 7, 13, and 10 factors), and the remaining factors were not analyzed. Survey cycles analyzed were 2008, 2010, 2012, 2014, 2016, 2018, 2020, 2021, and 2024 (2022 excluded). Mean importance ratings are available for all nine cycles unless noted. The percentage of program directors citing a factor is first reported for 2014, so at most six cycles (2014, 2016, 2018, 2020, 2021, 2024) are available for it; “ Years factors were ranked≥ 50%” counts the cycles in which the factor was cited by at least 50% of program directors, out of the cycles with a percentage available. “≥ 50% in 2024” is shown for information; it is not a statement of which factors entered the ≥ 50%-cited models. “Any failed attempt in USMLE” (surveyed from 2016) was excluded from the Theme 5 models. Factors surveyed in fewer than nine cycles (any failed attempt in USMLE; evidence of professionalism and ethics, IM; diversity characteristics, IM; history of overcoming significant obstacles, RAD) make those panels unbalanced. Spelling and capitalization of factor names were harmonized from the extraction file and should be checked against the NRMP reports. Spec. = specialty; ANES = anesthesiology; GS = general surgery; IM = internal medicine; RAD = radiology; MSPE = medical student performance evaluation; N = number of cycles with a percentage available. Each factor contributes one observation for every cycle with a mean rating (nine for factors available in all cycles); these observations sum to the 3-item counts in Supplementary Table 2.

**Supplementary Table 2**. **Factors entering the ≥ 50%-cited theme-level models, by specialty and theme (themes for which factor-level results were generated).**

| **Specialty** | **Theme** | **Factor (as extracted)** | **Survey years in ≥ 50%-cited model (of 6)** |
| --- | --- | --- | --- |
| Anesthesia | Education and Performance | AOA | 2 |
| Anesthesia | Education and Performance | Class ranking/quartile | 6 |
| Anesthesia | Education and Performance | Grades required in clerkships | 4 |
| Anesthesia | Education and Performance | Graduate of US allopathic medical school | 1 |
| Anesthesia | Education and Performance | Honors in clinical clerkships | 1 |
| Anesthesia | Education and Performance | MSPE | 6 |
| Anesthesia | Education and Performance | graduate of highly-regarded US medical school | 1 |
| Anesthesia | Interview Day | Feedback from current residents | 6 |
| Anesthesia | Interview Day | Interaction with faculty during interview and visit | 6 |
| Anesthesia | Interview Day | Interpersonal Skills | 6 |
| Anesthesia | Interview Day | interaction with house staff during interview and visit | 6 |
| Anesthesia | Other Knowledge of Applicant | Audition elective/rotation within your department | 5 |
| Anesthesia | Other Knowledge of Applicant | Letters of Recommendation | 6 |
| Anesthesia | Other Knowledge of Applicant | Personal prior knowledge of the applicant | 6 |
| Anesthesia | Personal Qualities | Consistency of grades | 3 |
| Anesthesia | Personal Qualities | Leadership qualities | 6 |
| Anesthesia | Personal Qualities | Percieved Commitment to Specialty | 5 |
| Anesthesia | Personal Qualities | Percieved interest in program | 4 |
| Anesthesia | Personal Qualities | Personal Statement | 4 |
| Anesthesia | Personal Qualities | Professionalism and ethics | 6 |
| Anesthesia | Personal Qualities | diversity characteristics | 1 |
| Anesthesia | Personal Qualities | history of overcoming significant obstacles | 2 |
| Anesthesia | Personal Qualities | other life experience | 2 |
| Anesthesia | Standard Licensure Test | Any failed attempt in USMLE | 2 |
| Anesthesia | Standard Licensure Test | USMLE Step 1 pass | 6 |
| Anesthesia | Standard Licensure Test | USMLE Step 2 | 6 |
| General Surgery | Education and Performance | AOA | 1 |
| General Surgery | Education and Performance | Class ranking/quartile | 5 |
| General Surgery | Education and Performance | Grades in clerkship in preferred speciaty | 3 |
| General Surgery | Education and Performance | Grades required in clerkships | 2 |
| General Surgery | Education and Performance | Graduate of US allopathic medical school | 1 |
| General Surgery | Education and Performance | MSPE | 6 |
| General Surgery | Interview Day | Feedback from current residents | 6 |
| General Surgery | Interview Day | Interaction with faculty during interview and visit | 6 |
| General Surgery | Interview Day | Interpersonal Skills | 6 |
| General Surgery | Interview Day | interaction with house staff during interview and visit | 6 |
| General Surgery | Other Knowledge of Applicant | Audition elective/rotation within your department | 1 |
| General Surgery | Other Knowledge of Applicant | Letters of Recommendation | 6 |
| General Surgery | Other Knowledge of Applicant | Personal prior knowledge of the applicant | 6 |
| General Surgery | Personal Qualities | Leadership Qualities | 6 |
| General Surgery | Personal Qualities | Percieved Commitment to Specialty | 6 |
| General Surgery | Personal Qualities | Percieved interest in program | 5 |
| General Surgery | Personal Qualities | Professionalism and ethics | 5 |
| General Surgery | Personal Qualities | diversity characteristics | 2 |
| General Surgery | Personal Qualities | history of overcoming significant obstacles | 2 |
| General Surgery | Personal Qualities | other life experience | 3 |
| General Surgery | Personal Qualities | personal statement | 5 |
| General Surgery | Standard Licensure Test | USMLE Step 1 pass | 5 |
| General Surgery | Standard Licensure Test | USMLE Step 2 | 6 |
| Internal Medicine | Education and Performance | AOA | 1 |
| Internal Medicine | Education and Performance | Class ranking/quartile | 6 |
| Internal Medicine | Education and Performance | Continous medical education w/o gaps | 3 |
| Internal Medicine | Education and Performance | Grades in clerkship in preferred speciaty | 4 |
| Internal Medicine | Education and Performance | Grades required in clerkships | 4 |
| Internal Medicine | Education and Performance | Honors in clerkship in desired specailty | 1 |
| Internal Medicine | Education and Performance | Honors in clinical clerkships | 1 |
| Internal Medicine | Education and Performance | MSPE | 6 |
| Internal Medicine | Education and Performance | graduate of US allopathic medical school | 1 |
| Internal Medicine | Education and Performance | lacks of gaps in medical education | 1 |
| Internal Medicine | Interview Day | Feedback from current residents | 6 |
| Internal Medicine | Interview Day | Interaction with faculty during interview and visit | 6 |
| Internal Medicine | Interview Day | Interpersonal skills | 6 |
| Internal Medicine | Interview Day | interaction with house staff during interview and visit | 6 |
| Internal Medicine | Other Knowledge of Applicant | Audition elective/rotation within your department | 1 |
| Internal Medicine | Other Knowledge of Applicant | Letters of Recommendation | 6 |
| Internal Medicine | Other Knowledge of Applicant | Personal prior knowledge of the applicant | 4 |
| Internal Medicine | Personal Qualities | Consistency of grades | 1 |
| Internal Medicine | Personal Qualities | Leadership Qualities | 5 |
| Internal Medicine | Personal Qualities | Percieved Commitment to Specialty | 4 |
| Internal Medicine | Personal Qualities | Percieved interest in program | 5 |
| Internal Medicine | Personal Qualities | Professionalism and ethics | 6 |
| Internal Medicine | Personal Qualities | diversity characteristics | 2 |
| Internal Medicine | Personal Qualities | geographic preferences | 1 |
| Internal Medicine | Personal Qualities | history of overcoming significant obstacles | 3 |
| Internal Medicine | Personal Qualities | other life experience | 2 |
| Internal Medicine | Personal Qualities | personal statement | 5 |
| Internal Medicine | Standard Licensure Test | Any failed attempt in USMLE | 5 |
| Internal Medicine | Standard Licensure Test | USMLE Step 1 pass | 6 |
| Internal Medicine | Standard Licensure Test | USMLE Step 2 | 6 |
| Radiology-Diagnostic | Education and Performance | AOA | 3 |
| Radiology-Diagnostic | Education and Performance | Class ranking/quartile | 6 |
| Radiology-Diagnostic | Education and Performance | Grades required in clerkships | 6 |
| Radiology-Diagnostic | Education and Performance | Honors in clinical clerkships | 1 |
| Radiology-Diagnostic | Education and Performance | MSPE | 6 |
| Radiology-Diagnostic | Education and Performance | graduate of US allopathic medical school | 1 |
| Radiology-Diagnostic | Education and Performance | graduate of highly-regarded US medical school | 1 |
| Radiology-Diagnostic | Interview Day | Feedback from current residents | 6 |
| Radiology-Diagnostic | Interview Day | Interaction with faculty during interview and visit | 6 |
| Radiology-Diagnostic | Interview Day | Interpersonal skills | 6 |
| Radiology-Diagnostic | Interview Day | interaction with house staff during interview and visit | 6 |
| Radiology-Diagnostic | Other Knowledge of Applicant | Audition elective/rotation within your department | 2 |
| Radiology-Diagnostic | Other Knowledge of Applicant | Letters of Recommendation | 5 |
| Radiology-Diagnostic | Other Knowledge of Applicant | Personal prior knowledge of the applicant | 5 |
| Radiology-Diagnostic | Personal Qualities | Consistency of grades | 3 |
| Radiology-Diagnostic | Personal Qualities | Leadership Qualities | 4 |
| Radiology-Diagnostic | Personal Qualities | Percieved Commitment to Specialty | 6 |
| Radiology-Diagnostic | Personal Qualities | Percieved interest in program | 5 |
| Radiology-Diagnostic | Personal Qualities | Professionalism and ethics | 6 |
| Radiology-Diagnostic | Personal Qualities | diversity characteristics | 1 |
| Radiology-Diagnostic | Personal Qualities | history of overcoming significant obstacles | 2 |
| Radiology-Diagnostic | Personal Qualities | other life experience | 1 |
| Radiology-Diagnostic | Personal Qualities | personal statement | 5 |
| Radiology-Diagnostic | Standard Licensure Test | Any failed attempt in USMLE | 1 |
| Radiology-Diagnostic | Standard Licensure Test | USMLE Step 1 pass | 6 |
| Radiology-Diagnostic | Standard Licensure Test | USMLE Step 2 | 6 |

Supplementary Table 2: Each factor entered a specialty-year observation in each survey year in which at least 50% of that specialty’s program directors cited it; the threshold was applied separately in each of the six survey years (2014, 2016, 2018, 2020, 2021, 2024), so factors may enter or leave across years. The last column gives the number of those years in which the factor qualified. Factors listed outside the 3-item set in Supplementary Table 3 are included here.

**Supplementary Table 3. Pooled interview-day models: factors, observations, and estimates.**

| **Specialty** | **Interview-day factor** | **3-item model: survey cycles** | **≥ 50%-cited model: survey years** |
| --- | --- | --- | --- |
| ANES | Interpersonal skills | 9 | 6 |
| ANES | Interaction with faculty during interview and visit | 9 | 6 |
| ANES | Feedback from current residents | 9 | 6 |
| ANES | Interaction with house staff during interview and visit | — | 6 |
| GS | Interpersonal skills | 9 | 6 |
| GS | Interaction with faculty during interview and visit | 9 | 6 |
| GS | Feedback from current residents | 9 | 6 |
| GS | Interaction with house staff during interview and visit | — | 6 |
| IM | Interpersonal skills | 9 | 6 |
| IM | Interaction with faculty during interview and visit | 9 | 6 |
| IM | Interaction with house staff during interview and visit | 9 | 6 |
| IM | Feedback from current residents | — | 6 |
| RAD | Interpersonal skills | 9 | 6 |
| RAD | Interaction with faculty during interview and visit | 9 | 6 |
| RAD | Interaction with house staff during interview and visit | 9 | 6 |
| RAD | Feedback from current residents | — | 6 |

Supplementary Table 3: For each specialty, the table lists the interview-day factors included in the pooled models and the number of survey cycles (3-item model) or survey years (≥50%-cited model) contributing data for each factor. A dash (—) indicates that the factor was not included in that model. ANES, anesthesiology; GS, general surgery; IM, internal medicine; RAD, radiology.

**Supplementary Table 4. Number of selection factors (clusters) and observations in each theme-level model.**

| **Specialty** | **Theme** | **3-item model** | | | | **≥ 50%-cited model** | | |
| --- | --- | --- | --- | --- | --- | --- | --- | --- |
|  |  | **Factors analyzed (see Supplementary Table 1)** | **n** | **Obs** | **Min p** | **n** | **Obs** | **Min p** |
| ANES | Education & Performance | MSPE; Class ranking/quartile; Grades required in clerkships | 3 | 27 | .009 | 7 | 21 | < .001 |
| ANES | Interview Day | Interpersonal skills; Interaction with faculty during interview and visit; Feedback from current residents | 3 | 27 | .009 | 4 | 24 | .0015 |
| ANES | Other Knowledge of Applicant | Personal prior knowledge of the applicant; Letters of recommendation; Audition elective/rotation within your department | 3 | 27 | .009 | 3 | 17 | .009 |
| ANES | Personal Qualities | Personal statement; Leadership qualities; Perceived commitment to specialty | 3 | 27 | .009 | 9 | 33 | < .001 |
| ANES | Standard Licensure Test | USMLE Step 2; USMLE Step 1 pass | 2 | 18 | .056 | 3 | 14 | .009 |
| GS | Education & Performance | MSPE; Class ranking/quartile; Grades required in clerkships | 3 | 27 | .009 | 6 | 18 | < .001 |
| GS | Interview Day | Interpersonal skills; Interaction with faculty during interview and visit; Feedback from current residents | 3 | 27 | .009 | 4 | 24 | .0015 |
| GS | Other Knowledge of Applicant | Personal prior knowledge of the applicant; Letters of recommendation; Audition elective/rotation within your department | 3 | 27 | .009 | 3 | 13 | .009 |
| GS | Personal Qualities | Personal statement; Leadership qualities; Perceived commitment to specialty | 3 | 27 | .009 | 8 | 34 | < .001 |
| GS | Standard Licensure Test | USMLE Step 2; USMLE Step 1 pass | 2 | 18 | .056 | 2 | 11 | .056 |
| IM | Education & Performance | MSPE; Class ranking/quartile; Grades required in clerkships | 3 | 27 | .009 | 10 | 28 | < .001 |
| IM | Interview Day | Interpersonal skills; Interaction with faculty during interview and visit; Interaction with house staff during interview and visit | 3 | 27 | .009 | 4 | 24 | .0015 |
| IM | Other Knowledge of Applicant | Personal prior knowledge of the applicant; Letters of recommendation; Audition elective/rotation within your department | 3 | 27 | .009 | 3 | 11 | .009 |
| IM | Personal Qualities | Diversity characteristics; Evidence of professionalism and ethics; History of overcoming significant obstacles | 3 | 19 | .009 | 10 | 34 | < .001 |
| IM | Standard Licensure Test | USMLE Step 2; USMLE Step 1 pass | 2 | 18 | .056 | 3 | 17 | .009 |
| RAD | Education & Performance | MSPE; Class ranking/quartile; Grades required in clerkships | 3 | 27 | .009 | 7 | 24 | < .001 |
| RAD | Interview Day | Interpersonal skills; Interaction with faculty during interview and visit; Interaction with house staff during interview and visit | 3 | 27 | .009 | 4 | 24 | .0015 |
| RAD | Other Knowledge of Applicant | Personal prior knowledge of the applicant; Letters of recommendation; Audition elective/rotation within your department | 3 | 27 | .009 | 3 | 12 | .009 |
| RAD | Personal Qualities | Personal statement; Perceived commitment to specialty; History of overcoming significant obstacles | 3 | 20 | .009 | 9 | 33 | < .001 |
| RAD | Standard Licensure Test | USMLE Step 2; USMLE Step 1 pass | 2 | 18 | .056 | 3 | 13 | .009 |

Supplementary Table 4: Each observation is one selection factor’s mean importance rating (1–5) in one survey cycle in one specialty; clusters are the selection factors. 3-item model: the top three factors per theme from the 2024 report (two for Standard Licensure Test, which excludes “any failed attempt in USMLE”), named in the table and described in Supplementary Table 3, across the nine survey cycles 2008–2024 (excluding 2022), unbalanced where a factor was not surveyed in every cycle. For example, ANES Education & Performance comprises 3 factors × 9 cycles = 27 observations. ≥ 50%-cited model: factors cited by at least 50% of the specialty’s program directors in a given survey year (2014–2024); the factors are listed in Supplementary Tables 1 and 2, and observations equal the sum over factors of the years in which each qualified. n = number of factors (clusters); Obs = number of factor-by-survey-cycle observations; Min p = smallest attainable wild cluster bootstrap p given the number of clusters (see Supplementary Methods). ANES = anesthesiology; GS = general surgery; IM = internal medicine; RAD = radiology.

**Supplementary Table 5. Model estimates**

| **Model** | **Clusters / observations** | **Common slope (95% CI)** | **Wild bootstrap 95% CI; p** | **Specialty × time: Wald F; cluster-robust p; bootstrap p** | **Level change at 2021 (95% CI)** | **Slope change after 2021 (95% CI); p** |
| --- | --- | --- | --- | --- | --- | --- |
| 3-item (per survey cycle) | 12 / 108 | 0.061 (0.051, 0.070) | 0.052, 0.069; p < .001 | F(3, 11) = 7.76; p = .005; p = .070 | −0.26 (−0.32, −0.20) | −0.06 (−0.15, 0.02); p = .14 |
| ≥ 50%-cited (per calendar year) | 16 / 96 | 0.005 (0.001, 0.009) | 0.0015, 0.0084; p = .007 | F(3, 15) = 1.06; p = .39; p = .46 | −0.09 (−0.14, −0.05) | −0.009 (−0.033, 0.014); p = .40 |

Supplementary Table 5: Cluster = factor within specialty (12 clusters × 9 survey cycles = 108 observations in the 3-item model; 16 clusters × 6 survey years = 96 observations in the ≥ 50%-cited model). The model includes a fixed effect for each cluster and a linear time trend; it does not include survey-wave fixed effects or two-way clustering, so correlation among factors from different specialties in the same wave is not modeled. Slopes are per survey cycle (3-item) or per calendar year (≥ 50%). Confidence intervals in the “Common slope” column are cluster-robust; the segmented model adds a post-2021 level change and slope change. CI = confidence interval; Wald F tests the specialty-by-time interaction.

**Supplementary Table 6. Variance inflation factors of the five theme composites.**

| **Theme** | **Raw VIF** | **Adjusted VIF** |
| --- | --- | --- |
| Theme 1: Education & Performance | 1.34 | 1.58 |
| Theme 2: Interview Day | 2.47 | 1.44 |
| Theme 3: Other Knowledge of Applicant | 3.25 | 1.42 |
| Theme 4: Personal Qualities | 2.64 | 1.49 |
| Theme 5: Standard Licensure Test | 1.87 | 1.16 |

Each composite is the mean rating of the theme’s items within a specialty-year (n = 36). Theme 5 excludes the “any failed USMLE attempt” item. Raw VIFs use the composites as constructed; adjusted VIFs use composites after removing specialty means and a linear survey-year trend. Raw pairwise correlations ranged from r = 0.13 to 0.75 (mean VIF 2.56); residual correlations ranged from r = 0.25 to 0.51 (mean VIF 1.38). VIF = variance inflation factor.

**Supplementary Table 7: Trends in the importance of thematic domains, by specialty.**

| **Thematic domain** | **ANES** | | | | **GS** | | | | **IM** | | | | **RAD** | | | |
| --- | --- | --- | --- | --- | --- | --- | --- | --- | --- | --- | --- | --- | --- | --- | --- | --- |
|  | **Slope** | **Naive p** | **Boot p** | **q** | **Slope** | **Naive p** | **Boot p** | **q** | **Slope** | **Naive p** | **Boot p** | **q** | **Slope** | **Naive p** | **Boot p** | **q** |
| **Theme 1: Education & Performance** | −0.008 | .471 | .657 | .772 | −0.012 | .431 | .601 | .752 | 0.010 | .027 | .135 | .296 | 0.003 | .378 | .146 | .296 |
| **Theme 2: Interview Day** | 0.000 | .870 | .912 | .912 | 0.009 | .048 | .062 | .296 | 0.006 | .056 | .172 | .296 | 0.004 | .041 | .171 | .296 |
| **Theme 3: Other Knowledge of Applicant** | 0.012 | .133 | .368 | .567 | 0.001 | .961 | .860 | .905 | 0.011 | .304 | .601 | .752 | 0.020 | <.001 | .115 | .296 |
| **Theme 4: Personal Qualities** | 0.037 | <.001 | .032 | .296 | 0.034 | <.001 | .053 | .296 | 0.043 | .008 | .178 | .296 | 0.002 | .641 | .484 | .692 |
| **Theme 5: Standard Licensure Test** | 0.016 | .093 | .114 | .296 | −0.013 | .519 | .718 | .798 | 0.019 | .094 | .133 | .296 | 0.051 | <.001 | .106 | .296 |

Supplementary Table 7: With G = 2 or 3 clusters, the Webb distribution contains only 6^G (36 or 216) weight vectors, or 18 or 108 distinct absolute t statistics, so the smallest attainable two-sided bootstrap p is approximately .056 (2 clusters) or .009 (3 clusters) regardless of the number of replications. Under Benjamini-Hochberg correction across 20 tests, a 2-cluster theme therefore cannot be rejected at q < .05, and a 3-cluster theme can be rejected only if at least four tests reach this minimum. Because theme-level tests are limited in this way, we added a post hoc pooled interview-day model across specialties (12 items in the 3-item analysis and 16 in the ≥50% analysis; item fixed effects; standard errors clustered on item; Webb wild cluster bootstrap), which estimated a common slope, tested specialty-by-time interaction, and fit a segmented model for level and slope change at 2021. The overall trends in Table 1 were assessed with the same wild cluster bootstrap and Benjamini-Hochberg correction across eight specialty-by-model tests.
